# Use of the Pharmacy First service in England in the first 12 months: geographic variation and health system context

**DOI:** 10.64898/2026.06.18.26355952

**Authors:** Weiyao Meng, Kimberley Sonnex, Aysel Pehlivanli, Thomas Allen, Elizabeth Dolan, Rebecca Glover, James Goulding, Hannah Higgins, Nicholas Mays, Amelia Taylor, Tracey Thornley, Anthony J Avery

## Abstract

**Objectives:** The Pharmacy First (PF) service was introduced across England from 31 January 2024 to expand the clinical role of community pharmacies and improve access to primary care. This paper describes use of PF in its first 12 months, in terms of uptake, access routes, consultation outcomes, geographic variations, service costs and antimicrobial supply.

**Methods:** A descriptive analysis of all PF consultations submitted for payment to NHS Business Services Authority in England between 31 January 2024 and 31 January 2025. Pharmacy-level consultation data were linked to national data on population, location and pharmacy characteristics. PF use was examined using population-standardised consultation rates and consultations per pharmacy.

**Results:** During the first year of implementation, 2,205,731 PF consultations were recorded as delivered across 11,349 pharmacies, with payment of £123 million to pharmacies. Uptake increased steadily over time. Most consultations were for acute sore throat (33%) and uncomplicated urinary tract infection (27%), with corresponding antibiotics, phenoxymethylpenicillin and nitrofurantoin being the most supplied. Most people self-referred (74%) into the service, with 95% of consultations managed without onward referral.

Substantial geographic variation was observed. Northern regions had higher use based on the eligible population. The South East and Midlands had higher activity per pharmacy. London showed a distinct pattern, with higher self-referral into the service, lower medication supply and higher referral to other healthcare services. Higher consultation volume was weakly associated with pharmacy characteristics, including opening hours, pharmacy type and retail setting, and local context, in terms of socio-economic and geographic factors.

**Conclusions:** PF had immediate uptake and is operating primarily as a direct-access model for common acute conditions. Findings suggest that PF is contributing to improved access to care and may shift demand away from general practice. However, the service uptake appears to be shaped by geographic location, proximity to other healthcare services and pharmacy characteristics.

## 1 Background

The COVID-19 pandemic highlighted long-standing supply constraints in General Practitioner (GP)-led primary care services in England, with increased demand reflected in longer appointment waiting times and delayed referrals to specialist services^1, 2^. To alleviate this pressure, the NHS Primary Care Recovery Plan (2023) identified increasing the clinical role of community pharmacists as a strategic priority^2^. The Pharmacy First (PF) service was launched across England as part of this policy shift on January 31, 2024^3^. Since then, the service has enabled patients to access assessment and treatment through community pharmacies for seven specific clinical conditions under Patient Group Directions^4^, namely: acute otitis media, uncomplicated urinary tract infection (UTI), acute sore throat, sinusitis, impetigo, shingles, and infected insect bites. Access to the service includes both self-referral and referral from other parts of the health system, for example from GPs or NHS111 (an online and telephone advice/triage service), aiming to improve the timeliness of treatment and reduce GP workload, while continuing to support antimicrobial stewardship^5^. Pharmacies are reimbursed through a combination of fixed and activity-based payments, designed to incentivise participation while maintaining cost control.

Previous services in England for managing minor ailments through pharmacies were largely commissioned locally and fragmented, resulting in significant inequalities in geographical access^6^. While PF aims to move beyond this patchwork to provide a standardised approach, it also builds on previous national initiatives such as the NHS Community Pharmacist Consultation Service, which offered pharmaceutical advice for minor ailments or non-urgent medication requests in pharmacies to patients referred from NHS111 or GPs^7^. However, in PF, established clinical pathways can be completed in the pharmacy, and prescription-only medication, such as antibiotics, can be supplied when necessary. Evaluations of similar models conducted in other nations in the UK (Scotland and Wales) support the feasibility and potential systemic benefits of expanding the clinical role of pharmacists^8, 9^.

Evidence regarding PF’s implementation remains limited. Previous studies have mainly focused on aggregated consultation counts. It remains unclear how PF operates within regions, whether regional uptake varies, and how service use relates to local population and geographic context. Understanding these patterns is important for informing future service development.

### Research objectives

This analysis aims to describe the first 12 months of PF use in England by examining:

1. National uptake of the service and temporal trends in consultations,
2. Access routes and consultation outcomes within PF consultations,
3. Geographic variation in PF use across sub-Integrated Care Board (ICB) areas and NHS England regions,
4. How pharmacy characteristics and local contextual factors are associated with patterns of PF use,
5. NHS reimbursement cost for PF, including per pharmacy and total payments,
6. Antimicrobial supply patterns for the treatment of the seven conditions included in PF.

## 2 Method

This analysis forms part of the national evaluation of PF^5^. It uses routinely collected administrative data to examine the first 12 months of PF implementation in England (1 February 2024 to 31 January 2025). The analysis focuses on PF service uptake, access routes, consultation outcomes, geographic variation at both pharmacy and regional levels, pharmacy and area-level characteristics associated with use, NHS reimbursement costs and antimicrobial supply.

### 2.1 Pharmacy characteristics and contextual variables

Community pharmacy activity reflects internal organisational characteristics, the population served and the local healthcare context. This analysis covers both **internal organisational features** (i.e., pharmacy opening hours and pharmacy business type) and **contextual characteristics** (i.e., pharmacy-level deprivation, rural–urban classification, retail centre classification, and proximity to other healthcare facilities). These variables provide a proxy for organisational capacity and describing the local environment in which pharmacies operate but do not capture staff composition or turnover.

### 2.2 Data sources and linkage

This analysis focuses predominantly on PF consultation data obtained from the NHS Business Services Authority Open Data Portal^10^, which records monthly consultations submitted by community pharmacies for NHS payment. Additional anonymised summary data on consultation outcomes and access route were obtained by agreement with NHS England via a Data Protection Impact Assessment and consultation totals differ slightly between the two sources due to differences in recording processes. Antimicrobial supply data were obtained from the NHSBSA ePACT2 PF prescribing dashboard^11^.

Pharmacies were linked via postcode to Lower Super Output Areas (LSOAs), Sub-ICB areas, ICBs, and NHS England regions using national lookup tables. Detailed data sources and full variable definitions are provided in Appendix Tables A1 and A2.

### 2.3 Analytical approach

To capture different dimensions of service activity, two use metrics were constructed:

1. **Consultations per 1,000 eligible population**, representing population-adjusted uptake and approximating patient demand for the service.
2. **Consultations per pharmacy**, representing pharmacy-level delivery intensity and provider engagement in service provision.

Population denominators are derived from Office for National Statistics mid-2024 population estimates at sub-ICB level^12^. The eligible population was constructed using condition-specific age-and sex-based eligibility criteria. More detailed clinical exclusion criteria within the PF pathways (e.g. pregnancy status or recurrent infection history) could not be operationalised at population level and were therefore not incorporated into denominator estimates.

Summary statistics (mean, median, standard deviation, interquartile range (IQR)) and inferential statistics (Spearman’s rank order correlation) were used.

### 2.4 PF cost and payment analysis

Pharmacies are reimbursed for providing PF services based on fixed and variable payments, consisting of:

1. An initial fixed payment of £2000 for each pharmacy that signed up to provide PF before 31st January 2024. This payment was recovered if the pharmacy delivered fewer than five PF consultations by 31st March 2024. The initial payment was to cover, for example, setting up new consultation room (if needed) for PF consultations and buying basic diagnostic equipment to be used in PF.
2. A monthly fixed payment of £1000 for each pharmacy that exceeds a minimum consultation threshold (details of thresholds in Appendix Table A3).
3. A variable payment of £15 per completed PF consultation below an upper consultation cap. From January 2024 to March 2024, there was no cap; from April 2024 to September 2024, there was a cap of 3000 consultations, and from October 2024 onwards, caps were based on the band the pharmacy was allocated (six possible bands), with the mean cap being 240 consultations.

Payments were computed by summing the initial fixed payment, monthly fixed payments, and consultation-based payments, with total payments defined as the sum of these components. Results are presented at both national level and per pharmacy to assess variation. Analyses also include the proportion of pharmacies qualifying for the monthly fixed payment by exceeding the minimum threshold, and the number of consultations that did not qualify for payment as they exceeded the consultation cap.

## 3 Results

### 3.1 National uptake and growth

#### 3.1.1 Overall service volume and growth

Between 1 February 2024 and 31 January 2025, 11,349 unique pharmacies delivered 2,205,731 consultations, after excluding four suspended pharmacies (15 consultations).

Monthly consultation volume (Appendix Table A5) increased from 125,270 consultations in February 2024 to 259,323 in December 2024, with a lower volume in January 2025. No evidence of plateauing was observed within the first 12 months.

#### 3.1.2 Distribution by clinical condition

Acute sore throat (33.4%) and uncomplicated UTI (27.5%) accounted for over 60% of all consultations; acute otitis media (11.8%), sinusitis (11.3%), infected insect bite (9.0%), impetigo (4.3%) and shingles (2.6%) composed the rest (Appendix Table A5).

Figure 1 suggests seasonal variation for acute sore throat, acute otitis media, and infected insect bite. Growth in total consultations over time was largely driven by the larger volumes of consultations recorded for acute sore throat and uncomplicated UTI.

**Figure 1:**
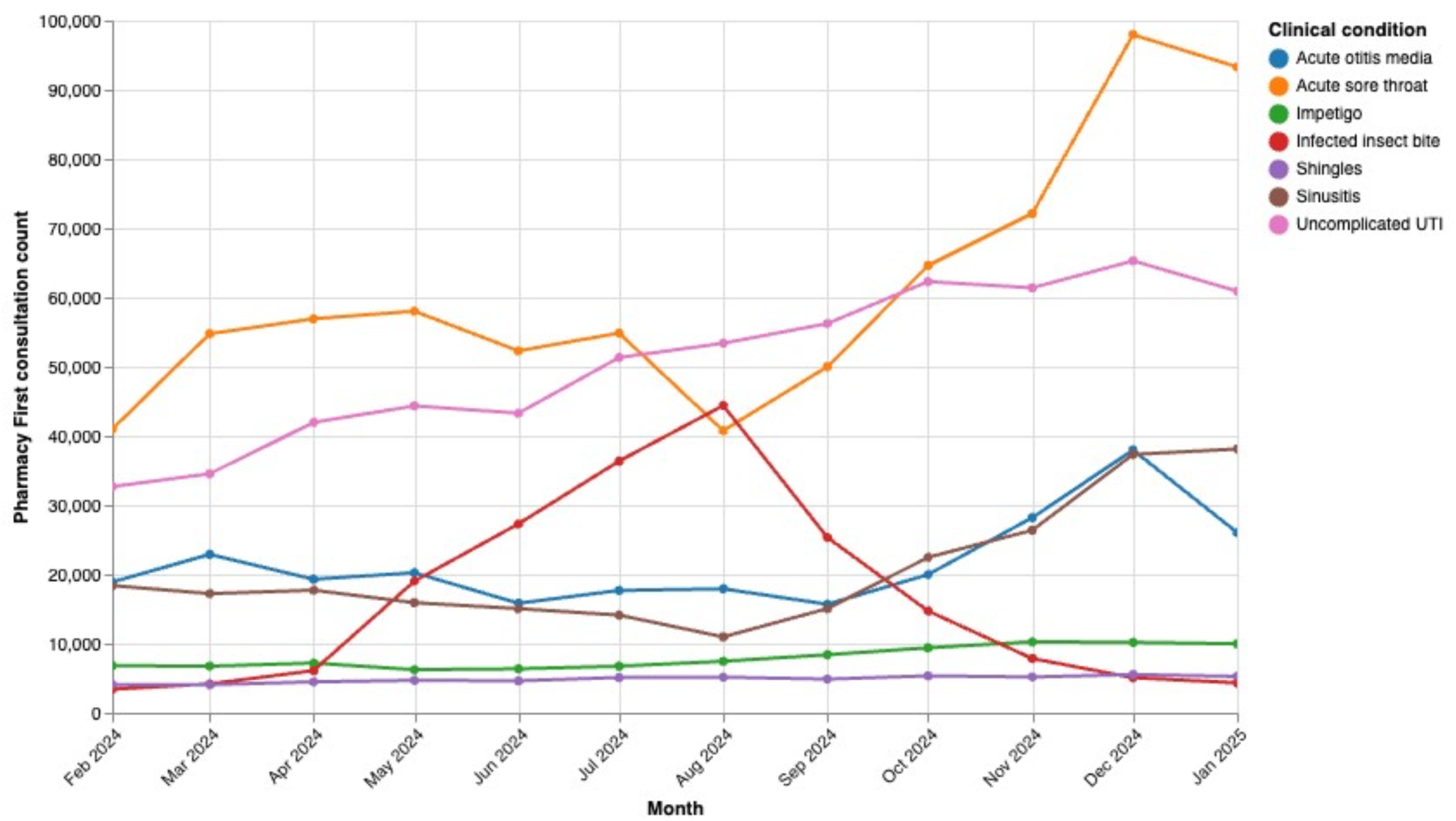
PF consultation trend over time for each clinical condition

### 3.2 Patient journeys: access routes and consultation outcomes

Most consultations were recorded as direct patient self-referral (74.2%), followed by GP referrals (12.1%) and signposting (an informal referral from another healthcare service; 11.0%) (Figure 2a). Referrals via NHS 111 accounted for 2.6% of consultations, while access from urgent or emergency care services was extremely rare (less than 0.1%).

**Figure 2:**
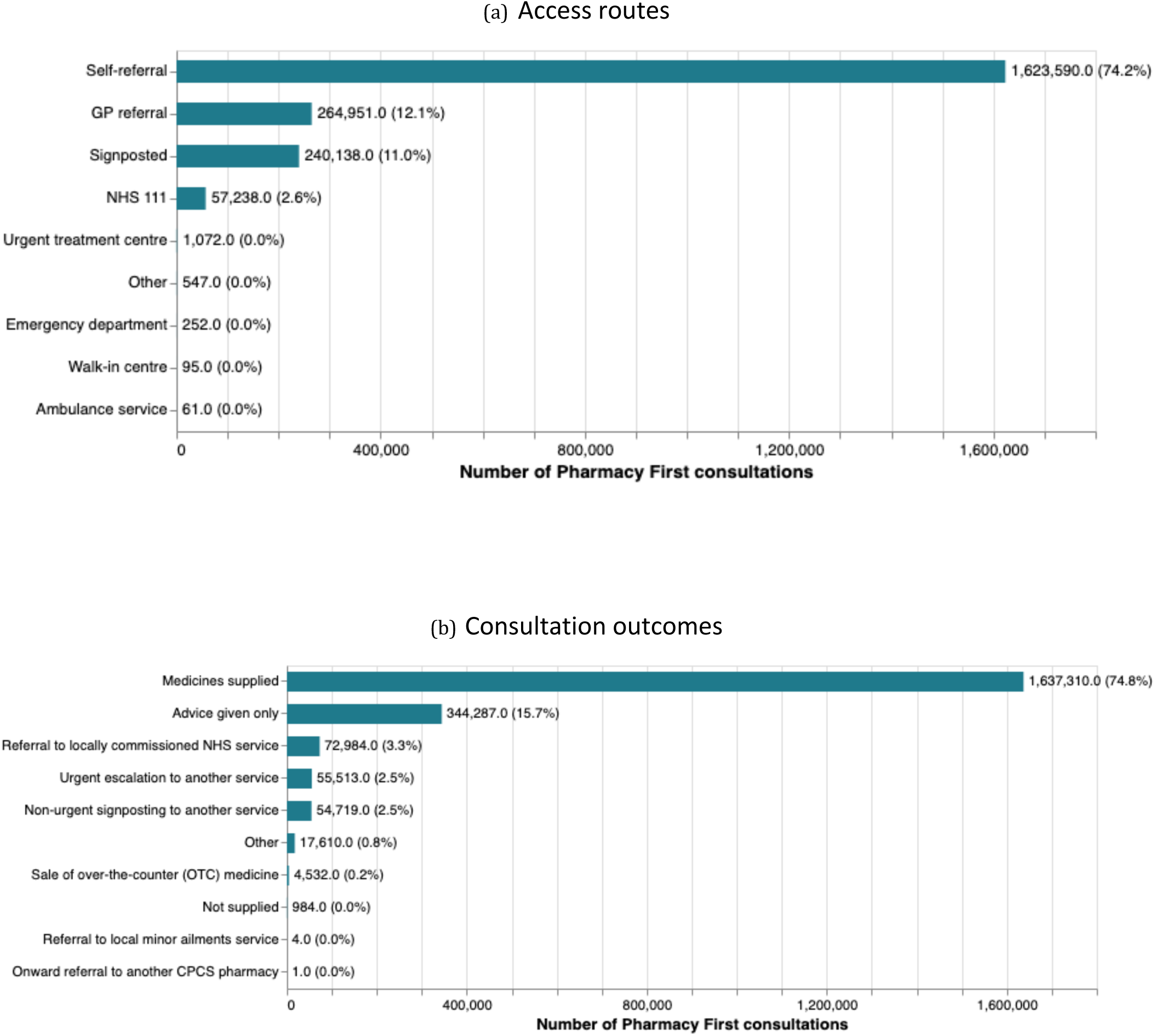
Distribution of PF consultations by (a) access route and (b) consultation outcome

Consultation outcomes demonstrate that almost 95% of PF consultations were managed without onward referral (Figure 2b), with 74.8% resulting in medicine supply and 15.7% recorded as “advice only”. Of the 5% of consultations requiring onward referral, half were classified as “urgent escalation to another service”.

The overall distribution of consultation access routes and outcomes remained broadly stable over time (Appendix Figure A1) and broadly similar across regions (Appendix Figure A2), with self-referral and in-pharmacy management predominating. London emerged as a clear regional outlier, demonstrating the highest proportion of self-referrals, a comparatively lower proportion of medicine supply, and higher proportions of referral to locally commissioned NHS services.

### 3.3 Geographic variation in Pharmacy First uptake

To ensure stable geographic linkage between use of PF by pharmacy, pharmacies with postcode changes during the study window were excluded. This excluded 37 pharmacies (0.3%) and 4,678 consultations (0.2% of total volume).

#### 3.3.1 Variation across Sub-ICBs

Substantial heterogeneity in PF utilisation was observed across sub-ICBs. The mean use rate was 40.0 consultations per 1,000 eligible population (median 38.8; IQR 35.6–44.8), ranging from 24.6 to 78.7 (Appendix Figure A3).

Monthly PF use increased across sub-ICBs, from a mean of 2.4 consultations per 1,000 eligible population (median 2.4; IQR 1.9-2.9) in February 2024 to 4.6 (median 4.5; IQR 4.1-5.0) in December 2024, remaining high at 4.2 (median 4.1; IQR 3.7-4.5) in January 2025 (Appendix Figure A4).

When expressed as consultations per pharmacy, the mean use rate across sub-ICBs was 198.9 consultations per pharmacy (median 195.1; IQR 174.5–220.6) during the first 12 months.

#### 3.3.2 Variation across NHS England regions

Across NHS England regions (Table 1), the two Northern regions tended to show higher use based on the eligible population, whereas London and the South East recorded lower rates. However, when expressed per pharmacy, the South East and Midlands showed higher activity per pharmacy, while London again was lowest.

**Table 1:** PF uptake across NHS England regions by total consultation count, per 1,000 eligible population, and per pharmacy.

| Region | Total volume | Per 1,000 eligible population | Per pharmacy | Eligible population |
| --- | --- | --- | --- | --- |
| North West | 317,595 | 43.25 | 190.86 | 7,342,556 |
| North East and Yorkshire | 364,096 | 41.98 | 190.63 | 8,672,712 |
| Midlands | 449,984 | 40.39 | 205.75 | 11,140,297 |
| South West | 207,249 | 35.44 | 200.43 | 5,848,629 |
| East of England | 241,191 | 35.36 | 204.23 | 6,821,747 |
| South East | 317,124 | 34.33 | 209.60 | 9,236,294 |
| London | 303,799 | 33.82 | 167.01 | 8,984,054 |

### 3.4 Pharmacy characteristics associated with PF consultations

#### 3.4.1 Internal organisational characteristics

Pharmacy opening hours were positively associated with PF consultation volume across all clinical conditions (Spearman’s *ρ* ranging from 0.2 to 0.4), suggesting that pharmacies with longer average weekly opening hours tended to deliver more consultations. The association was strongest for uncomplicated UTI.

PF uptake also differed by pharmacy type and region (Figure 3a). Nationally, most community pharmacies in PF are independent (57.8%), followed by high street chains (28.7%), other chains (7.5%) and supermarket pharmacies (6.0%). Independent pharmacies accounted for the largest share of consultations (56.3%) in each region and delivered substantially more consultations than other pharmacy types in London. Large supermarket pharmacies had higher median consultation volumes per pharmacy (median 286 consultations; range 133-1032), whereas independent pharmacies showed substantial heterogeneity (median 160 consultations; range 0–5489), with a small number of high-volume providers contributing disproportionately to overall activity.

**Figure 3:**
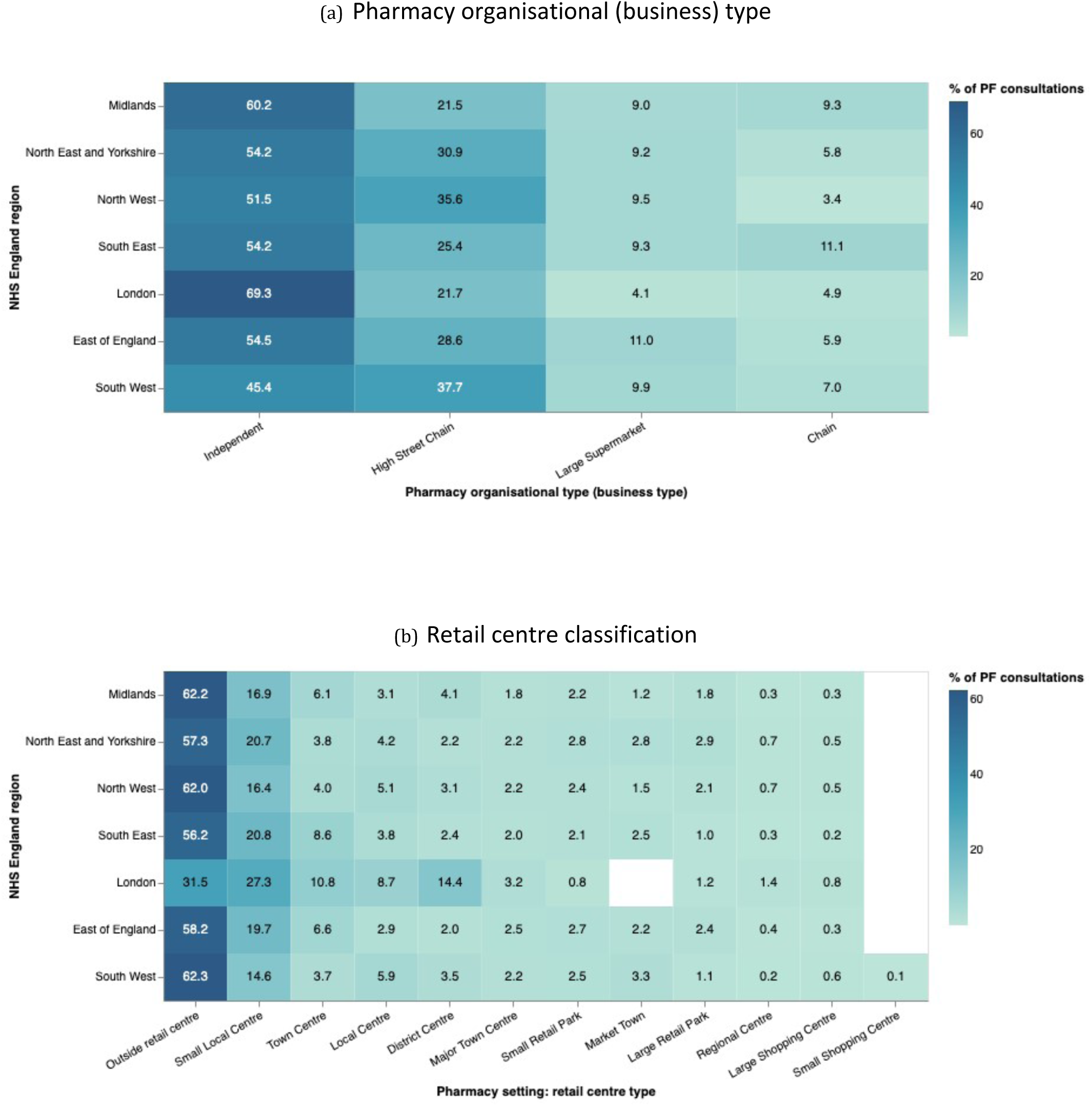
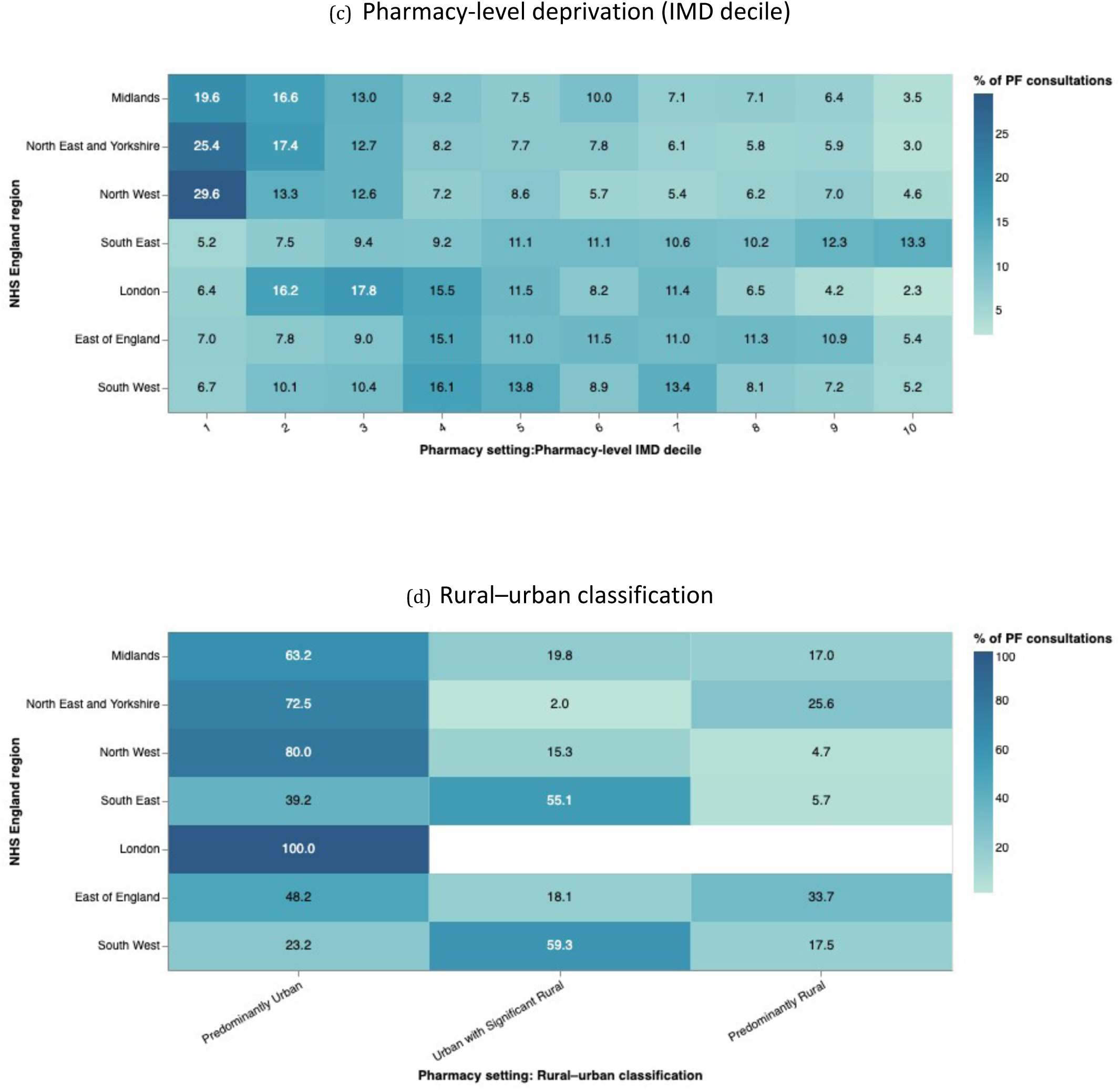
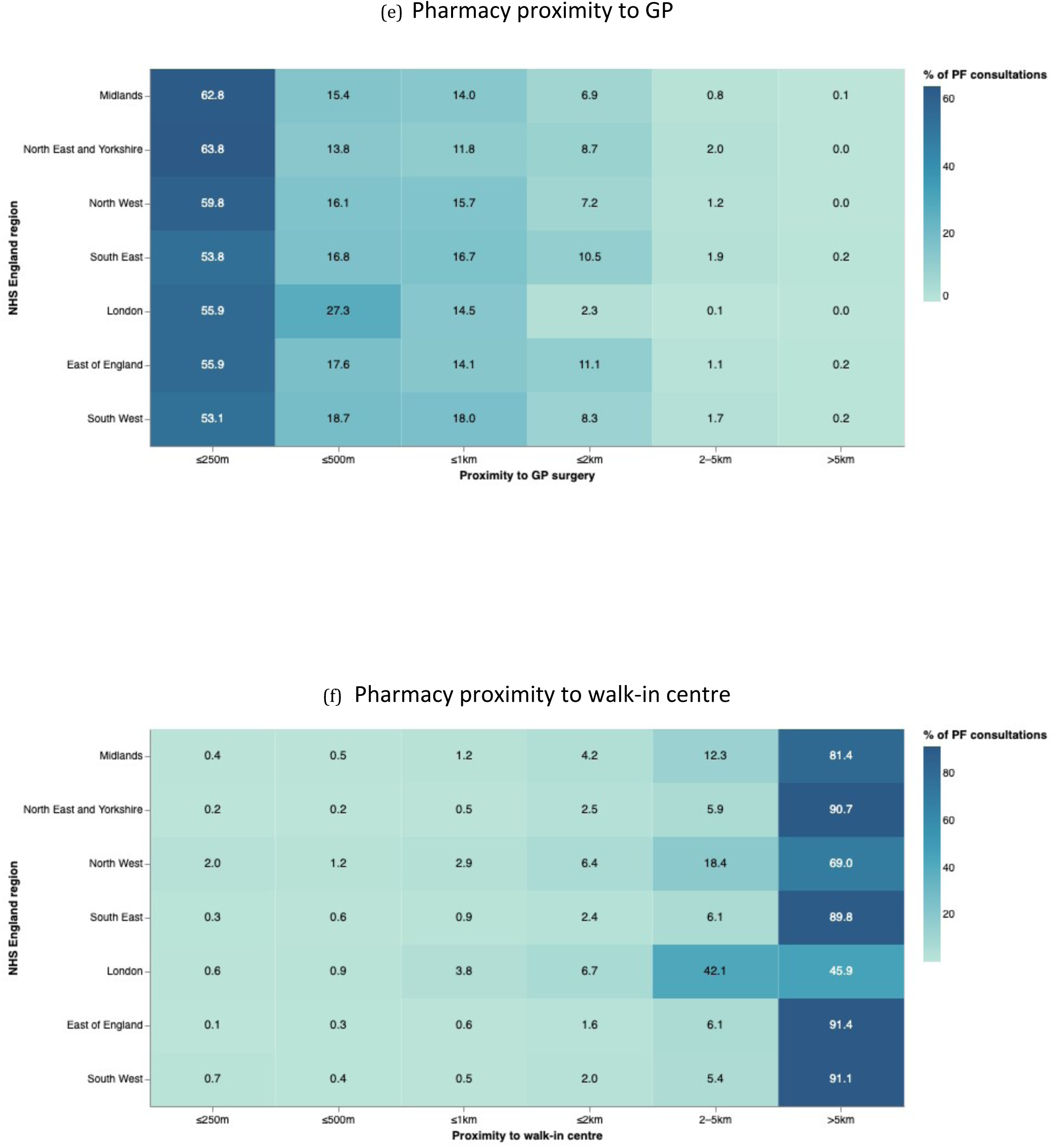
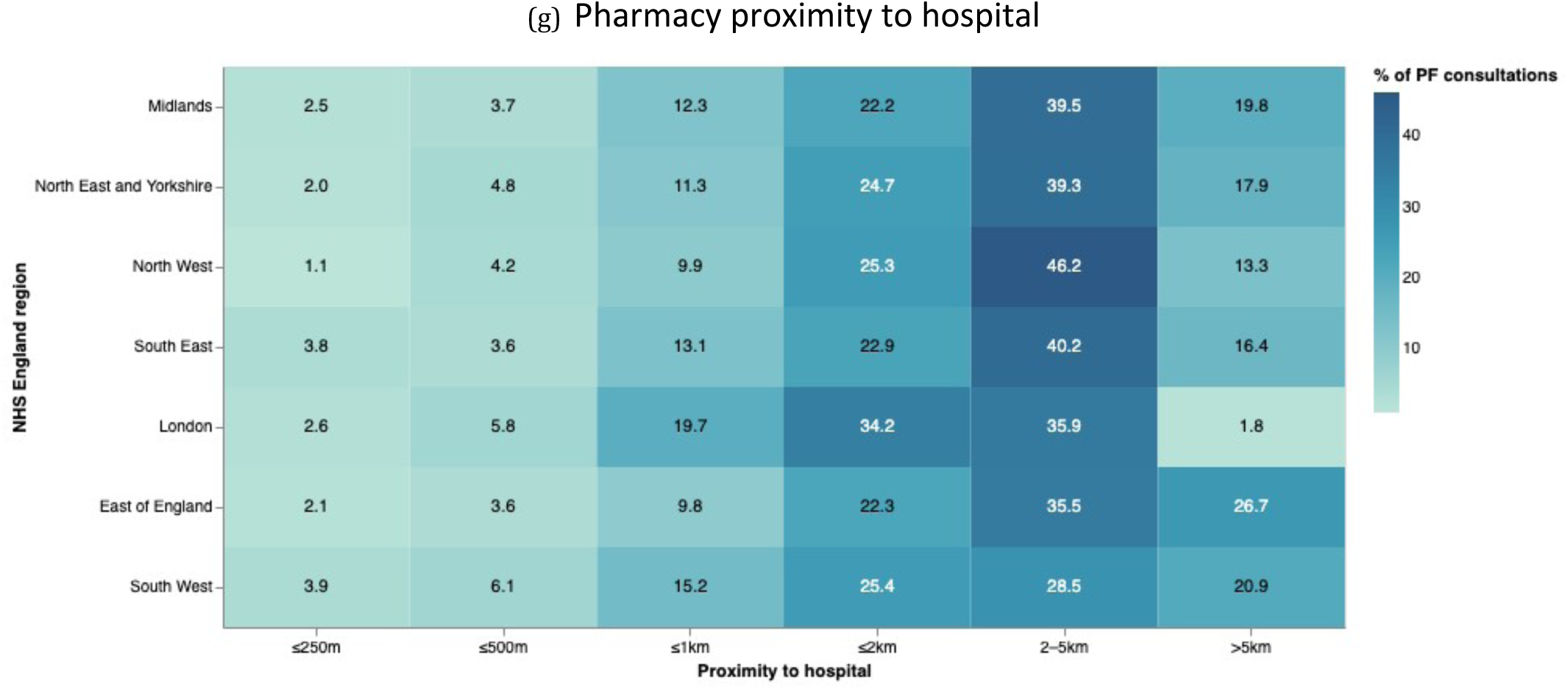
Regional distribution of PF consultations by pharmacy characteristics. Within each region, percentages represent the proportion of consultations accounted for by each category shown in each subplot.

#### 3.4.2 Pharmacy setting and accessibility

Regional comparisons further showed that local area contextual characteristics were associated with the distribution of PF consultations across pharmacies. In all regions, except London, pharmacies outside retail centres accounted for the largest proportion of PF consultations (approximately 56-62%; Figure 3b). London showed a more mixed distribution, with a higher share of consultations delivered by pharmacies located within central retail areas.

In Northern regions, most PF consultations were delivered by pharmacies located in predominantly urban areas and in more deprived neighbourhoods (Figure 3c-d). However, this association was less evident in Southern regions.

Accessibility to other healthcare services was also associated with consultation activity. Higher consultation volumes were observed among pharmacies located within walking distance of GP practices and among pharmacies located further from hospitals and walk-in centres (Figure 3e-g).

### 3.5 Cost and payment threshold patterns

Total NHS PF service payments to all pharmacies were £122,787,610. The initial £2000 payments were made to 9,701 pharmacies (£19,402,000), with a further 551 pharmacies receiving this payment only to have it recovered later for not meeting the initial consultation target. The fixed monthly payment of £1000 for exceeding the variable threshold (Appendix Table A3) was paid in 55.4% of the total possible months (£70,383,000). The consultation payments came to £33,002,610, with 864 consultations over the upper consultation cap and not qualifying for payment.

Many pharmacies did not meet the minimum consultation threshold and did not receive the £1000 monthly payment (Figure 4). As the threshold increased over time and became harder to meet, the percentage of pharmacies receiving the £1000 payment declined. Between February 2024 and September 2024, the threshold increased from 1 to 20 consultations per month, while the percentage of pharmacies receiving threshold payments decreased from 79% to 33%. Between September 2024 and December 2024, the threshold was held at 20, and the percentage receiving payment increased until January 2026, when the threshold was increased to 25.

**Figure 4:**
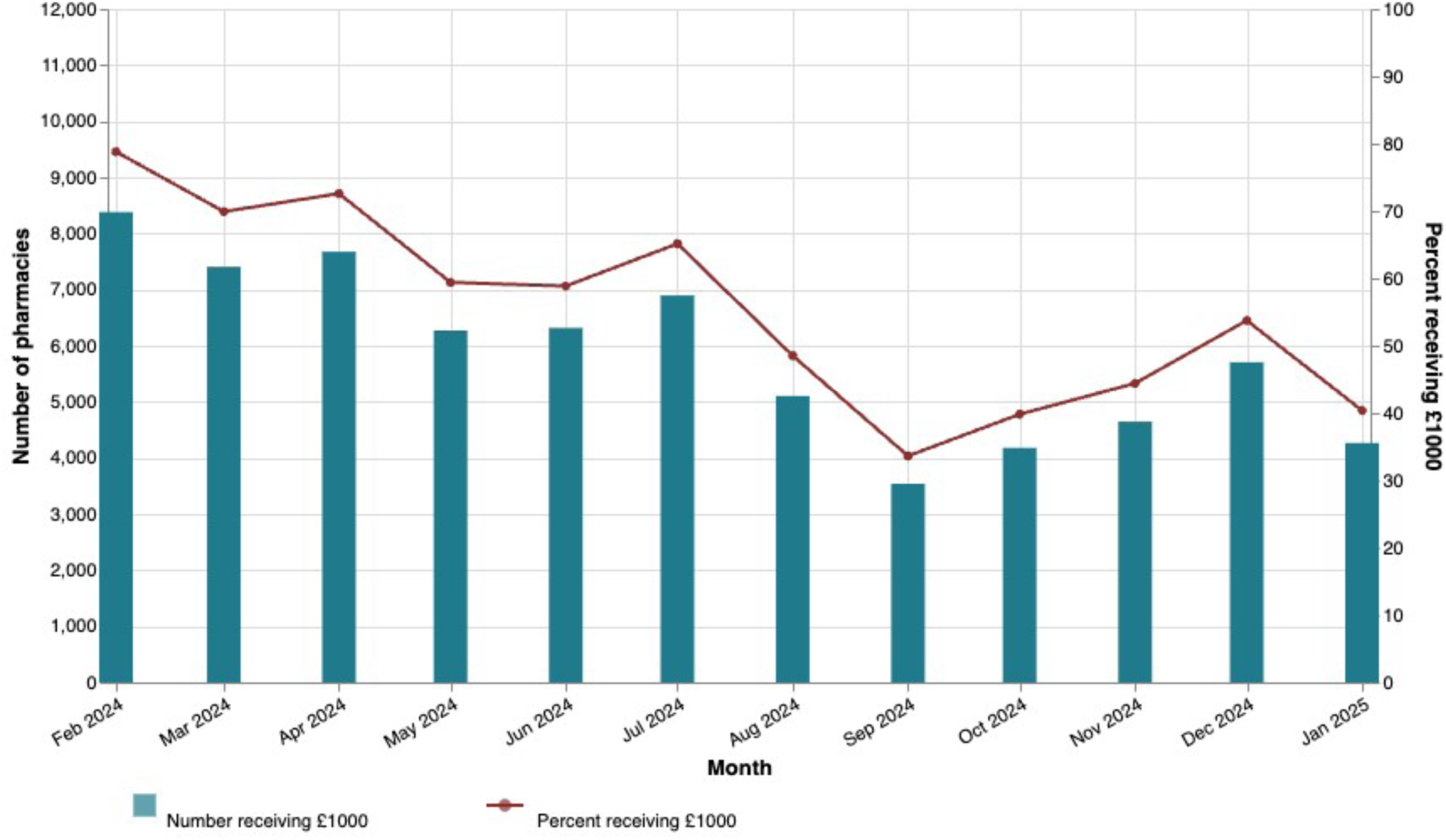
Pharmacies meeting minimum consultation threshold.

Over the first 12 months, pharmacies received a mean of £10,873 (median: £10,745; standard deviation: £6,771) in total payments, with 5% of pharmacies receiving over £20,900. There was little variation in total payments per pharmacy by NHS England region.

### 3.6 Supply of antimicrobials via Pharmacy First

A total of 1,524,327 antimicrobial items were supplied (Appendix Figure A5) across 2,205,731 consultations over the first 12 months, with 69.1 percent of consultations resulting in patients receiving an antimicrobial item. Phenoxymethylpenicillin (penicillin V) comprised the largest proportion of this supply (542,819 items; 35.6% of total supply), predominantly among patients on the acute sore throat and/or acute sinusitis pathways. This was followed by nitrofurantoin (513,432 items; 33.7% of total supply) and flucloxacillin (159,999 items; 10.5%). Total monthly supply peaked in December 2024 (173,037 items), and lowest in February 2024 (80,299 items), broadly mirroring consultation trends.

## 4 Discussion

### 4.1 Summary of empirical findings

PF use began immediately in its first year, with sustained growth and no clear plateau. Most consultations were self-referral and managed without onward referral. This indicates that PF is currently functioning as a first contact primary healthcare service. Some geographic and pharmacy level heterogeneity was observed, suggesting that PF is a uniform national policy in the design, but its uptake varies locally.

### 4.2 Pharmacy First’s uptake in the first year

The high initial uptake by pharmacies may have been associated with pre-existing minor ailment schemes^13^ and existing working relationships between pharmacies and general practice^14^. Therefore, pharmacies were extending capabilities that are already familiar to the public and general practice^14^. However, it was initially anticipated that PF, alongside other advanced services such as hypertension case finding, had the potential to free up 10 million GP appointments.

The absence of a clear plateau indicates that the service had not reached saturation, suggesting that public awareness and familiarity with the service were still developing. PF is one of an increasing number of advanced pharmacy services, including seasonal influenza vaccination and hypertension case finding^15^. In an increasingly financially pressured environment, community pharmacists argued that remuneration for these clinical services was insufficient and harmed their ability to deliver these additional clinical services. This may limit PF uptake^16^. Ongoing evaluation is needed as the service becomes more established.

### 4.3 Condition-specific patterns

High sore throat consultation volume warrants further investigation, particularly given recent evidence of relatively high antibiotic supply rates under the PF sore throat pathway in England compared with alternative models in Wales that incorporate point-of-care testing^17^.

Uncomplicated UTI consultations demonstrated a stronger association with longer pharmacy opening hours than other conditions. This may reflect the characteristics of the eligible population, which mainly consists of working-age women who are more likely to seek treatment in a convenient location and outside normal working hours. This suggests that PF use should be interpreted in the context of pathway-specific patient demand.

### 4.4 Pharmacy First operating as a context-embedded intervention

PF activity varied across regions and regional ordering shifted depending on whether uptake was measured per eligible population or per pharmacy. London demonstrated a distinct profile, characterised by higher self-referral, lower medicine supply, and greater onward referral to locally commissioned NHS services. This may reflect influence from population characteristics. Central London’s pharmacies may serve a relatively transient and younger working-age population^18^, which may influence condition severity and consultation outcomes. Also, London has a more complex healthcare infrastructure than other parts of the country, including more urgent care centres and other walk-in services. PF may therefore operate more frequently as a diversion or triage service.

Higher use was observed in pharmacies located further from hospitals or walk-in centres, showing substitution behaviour where alternative urgent care services are less accessible. However, PF use was also higher among pharmacies located close to GP practices. This may reflect the co-location of pharmacies within neighbourhood primary care hubs. These findings suggest that PF’s function may vary depending on the surrounding healthcare context, acting as a substitute, supplement, or triage service.

### 4.5 Pharmacy-level heterogeneity

While independent pharmacies contributed the largest share of total consultations, considerable heterogeneity was observed within this group, and large supermarket pharmacies had the highest per pharmacy consultation rate. Differences in incentive structures to deliver PF and staff composition, including the use of locum pharmacists and two-pharmacist models^14^, may influence engagement, which requires further evaluation.

### 4.6 Pharmacy First costs to the NHS

The community pharmacy sector has raised concerns about underfunding^19^ with schemes like PF offering much needed additional sources of income. PF payments totalled approximately £123 million over the first 12 months, representing close to 4.5% of the 2024/25 NHS funding for community pharmacy (£2.7 billion) and 57% of the £215 million additional funding allocated for PF^20^. That more of the PF funding allocation was not spent in the first 12 months may indicate that pharmacy uptake and patient demand fell below original expectations.

At pharmacy level, the almost £11,000 received on average likely represents a significant income boost. However, many pharmacies failed to provide sufficient consultations to qualify for the £1000 fixed monthly payment and missed out on this additional income. At the margin around these consultation thresholds, the impact of this missed payment is substantial. For example, in September 2024, when the threshold was set as 20 consultations, a pharmacy providing 19 consultations would receive £285 in payments, whereas a pharmacy providing 20 consultations would receive £1300. The additional consultation at the threshold has a marginal revenue of £1015, providing a large incentive to meet the threshold. How pharmacies respond to these incentives and how that response both changes over time and varies by pharmacy characteristics is an important area for future research.

### 4.7 Limitations

This study is limited to the reliance on routinely collected administrative data, with some variables reflecting pharmacy-recorded categories that may not fully capture clinical processes. For example, informal GP signposting may be recorded as self-referral if patients present without a formal electronic referral, potentially underestimating GP-mediated access.

In data processing, pharmacy business type was inferred using pharmacy names and may not fully reflect organisational diversity within national chains. Patient-level demographic and clinical information was unavailable, limiting investigation of equity, substitution behaviour, and populations served by PF. Population-standardised rates were therefore estimated using resident populations within administrative geographic boundaries rather than pharmacy catchment populations or commuter populations.

Finally, the analyses are descriptive and do not establish causal relationships. These limitations will be addressed in future mixed-method analyses within the wider evaluation programme^5^.

### 4.8 Policy implications

The steadily rising uptake observed during the first year of implementation suggests that nationally coordinated pharmacy-based services can be integrated quickly into existing primary care systems. The substantial consultation volume delivered within a short period indicates that community pharmacies can take on additional clinical responsibilities when supported by clear national clinical frameworks, whereas training and reimbursement mechanisms need to better support the provision of this service for reaching its full potential of freeing up 10 million GP appointments.

The findings emphasise the importance of local health system context in shaping service use. Variation in PF uptake across regions appears to reflect differences in population characteristics, pharmacy characteristics, and the availability and accessibility of alternative healthcare services. This suggests that the impact of nationally set PF service specifications will depend on the local systems in which they operate.

These insights have broader international relevance. Many health care systems are exploring UK pharmacy models to improve primary care access and reduce GP pressure, including Australia^21^, New Zealand^22^ and Canada^23^. The findings highlight the importance of considering local health system structure and complementary metrics (such as population-standardised utilisation and pharmacy workload) when implementing and assessing such services.

## 5 Conclusions

PF use increased rapidly during its first year of national implementation, delivering substantial consultation activity through community pharmacies. Most consultations were self-referrals, managed within pharmacies, and were concentrated in patients with acute sore throat and uncomplicated UTI, while uptake for other conditions remained comparatively low. These patterns suggest that PF is contributing to improved access to care and may be shifting demand away from GPs.

However, marked geographic and pharmacy-level variation was observed. Although PF is nationally standardised in design, use varied with factors such as pharmacy types, opening hours, local setting and accessibility. This suggests that PF might be used differently across areas, for example, as a direct treatment alternative to GP care in some areas, and more as a triage or referral service in others. As this study does not directly measure substitution away from GP, these interpretations remain indicative. Continued monitoring and linked analyses will be important to assess the longer-term impact of the service to GP services and urgent care demand.

## Data Availability

This study used a combination of publicly available datasets and restricted administrative data obtained under data sharing agreements. Publicly available data sources are referenced within the manuscript. Restricted data used in this study are not publicly available due to information governance and data sharing restrictions. Aggregated summary data may be available from the corresponding author upon reasonable request and subject to relevant approvals.

## Acknowledgements

We would like to thank members of the wider Pharmacy First Evaluation Collaboration for their contribution to the overall evaluation programme and ongoing discussions relating to interpretation of the findings.

## Ethics approval and informed consent statements

This study was given a favourable opinion by the University of Nottingham Faculty of Medicine and Health Sciences Research Ethics Committee, reference number FMHS 236-0724.

## Conflict of Interest Statement

Anthony J Avery has received funding from the NIHR, received support for attending meetings from NIHR, and has been National Clinical Director for Prescribing for NHS England since April 2022. All other authors declare no conflicts of interest.

## Funding statement

This study is funded by the NIHR Health and Social Care Delivery Research Programme, NIHR160217 Mixed-method impact and implementation evaluation of the “Pharmacy First” Services for the management of common conditions. The views expressed are those of the author(s) and not necessarily those of the NIHR or the Department of Health and Social Care.

## Appendix A. Tables and figures

**Table A1:** Data sources used in the study.

| Dataset | Provider | Level | Description |
| --- | --- | --- | --- |
| Pharmacy First consultation dataset<sup>10</sup> | NHS Business Services Authority (NHSBSA) | Pharmacy | Administrative dataset containing records of Pharmacy First consultations submitted for payment |
| Consolidated Pharmaceutical List (CPL)<sup>24</sup> | NHSBSA | Pharmacy | National register of community pharmacies in England |
| Population estimates (mid-2024)<sup>12</sup> | Office for National Statistics (ONS) | ICB | Mid-year population estimates by age and sex for health geographies |
| GP Workforce statistics<sup>25</sup> | NHS Digital | Practice | Monthly statistics describing GP workforce capacity |
| Index of Multiple Deprivation (IMD)<sup>26</sup> | ONS | LSOA | Area-level measure of socioeconomic deprivation |
| Rural–Urban Classification<sup>27</sup> | Department for Environment, Food & Rural Affairs (DEFRA), UK | Geographic lookup | Classification of areas according to rural and urban settlement patterns |
| Retail Centre Boundaries<sup>28</sup> | Geographic Data Service | Geographic | Dataset describing the spatial boundaries, catchment zones, local openness indicators, and retail typology of retail agglomerations across the UK. |
| Points of Interest (POI)<sup>29</sup> | Ordnance Survey | Location | Dataset containing locations of healthcare facilities and transport infrastructure |

**Table A2:**
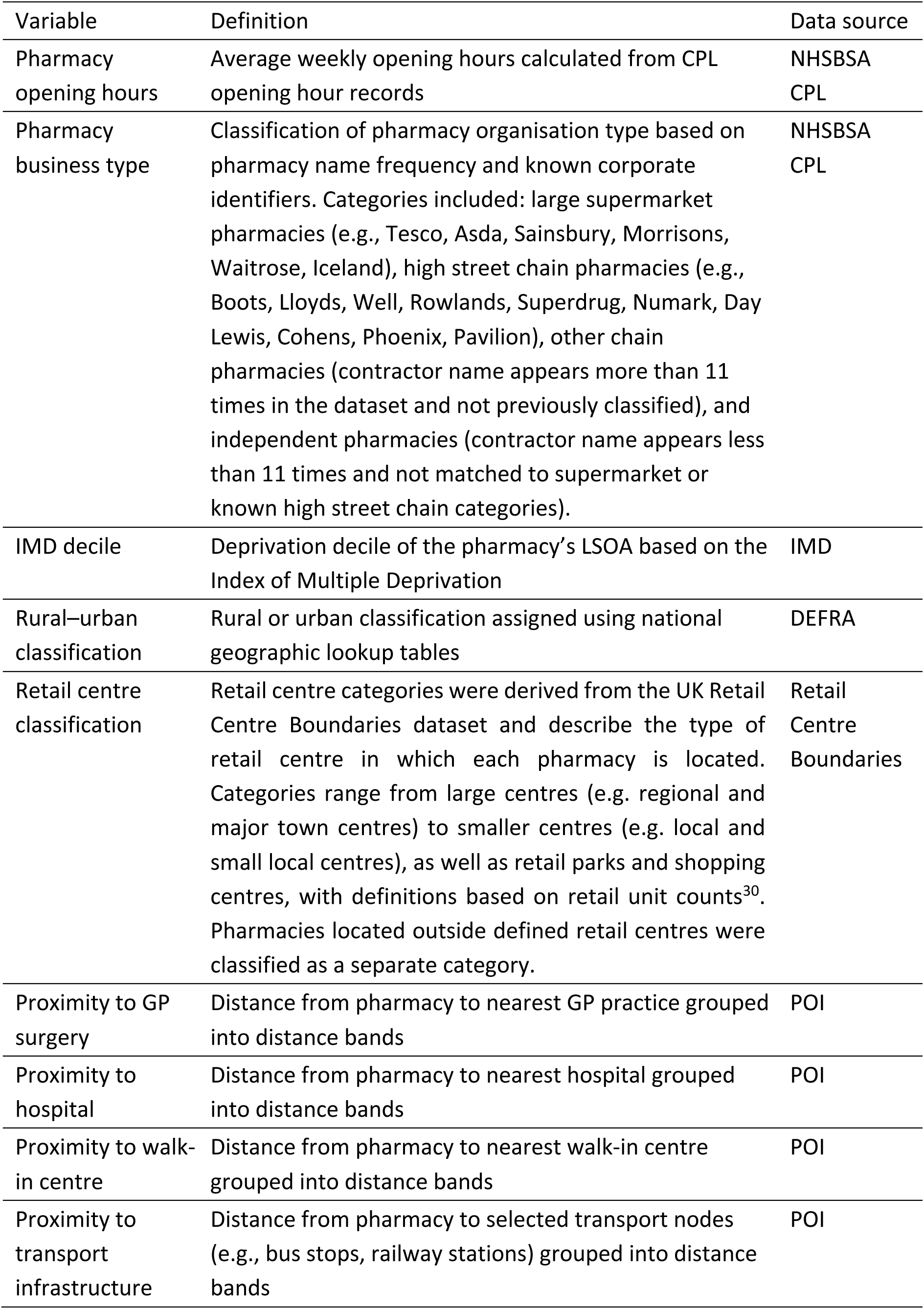
Pharmacy characteristic variables used in the analysis.

| Variable | Definition | Data source |
| --- | --- | --- |
| Pharmacy opening hours | Average weekly opening hours calculated from CPL opening hour records | NHSBSA<br>CPL |
| Pharmacy business type | Classification of pharmacy organisation type based on pharmacy name frequency and known corporate identifiers. Categories included: large supermarket pharmacies (e.g., Tesco, Asda, Sainsbury, Morrisons, Waitrose, Iceland), high street chain pharmacies (e.g., Boots, Lloyds, Well, Rowlands, Superdrug, Numark, Day Lewis, Cohens, Phoenix, Pavilion), other chain pharmacies (contractor name appears more than 11 times in the dataset and not previously classified), and independent pharmacies (contractor name appears less than 11 times and not matched to supermarket or known high street chain categories). | NHSBSA<br>CPL |
| IMD decile | Deprivation decile of the pharmacy's LSOA based on the Index of Multiple Deprivation | IMD |
| Rural–urban classification | Rural or urban classification assigned using national geographic lookup tables | DEFRA |
| Retail centre classification | Retail centre categories were derived from the UK Retail Centre Boundaries dataset and describe the type of retail centre in which each pharmacy is located. Categories range from large centres (e.g. regional and major town centres) to smaller centres (e.g. local and small local centres), as well as retail parks and shopping centres, with definitions based on retail unit counts<sup>30</sup>. Pharmacies located outside defined retail centres were classified as a separate category. | Retail Centre Boundaries |
| Proximity to GP surgery | Distance from pharmacy to nearest GP practice grouped into distance bands | POI |
| Proximity to hospital | Distance from pharmacy to nearest hospital grouped into distance bands | POI |
| Proximity to walk-in centre | Distance from pharmacy to nearest walk-in centre grouped into distance bands | POI |
| Proximity to transport infrastructure | Distance from pharmacy to selected transport nodes (e.g., bus stops, railway stations) grouped into distance bands | POI |

**Table A3:**
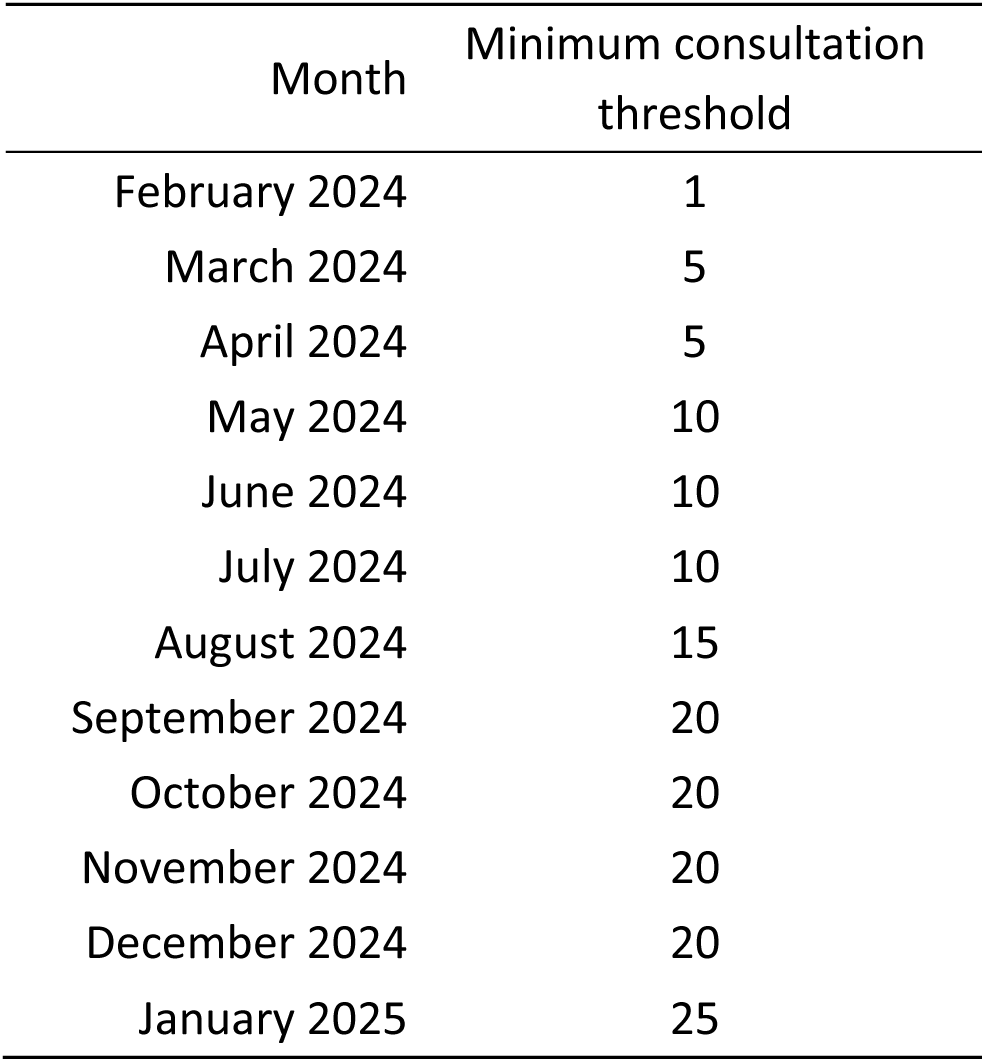
Monthly minimum consultation thresholds required to qualify for the fixed payment under the Pharmacy First service.

**Table A4:** Distribution of PF consultations by clinical condition for distance-selling pharmacies and all pharmacies.

|  | Acute sore throat | Uncomplicated UTI | Acute otitis media | Sinusitis |
| --- | --- | --- | --- | --- |
| DSP consultations (within-DSP %) | 6,552 (42.2%) | 4,438 (28.6%) | 179 (1.2%) | 2,389 (15.4%) |
| All consultations (share of total) | 736,638 (33.4%) | 607,644 (27.5%) | 260,589 (11.8%) | 248,701 (11.3%) |
| DSP share of all consultations | 0.89% | 0.73% | 0.07% | 0.96% |
|  | Infected insect bite | Impetigo | Shingles | Total |
| DSP consultations (within-DSP %) | 1,127 (7.3%) | 541 (3.5%) | 302 (1.9%) | 15,528 |
| All consultations (share of total) | 198,069 (9.0%) | 95,714 (4.3%) | 58,361 (2.6%) | 2,205,716 |
| DSP share of all consultations | 0.57% | 0.57% | 0.52% | 0.70% |

**Table A5:** Monthly Pharmacy First consultation counts by clinical conditions.

| Month | Acute<br>sore<br>throat | Uncomplicated<br>UTI | Acute<br>otitis<br>media | Sinusitis | Infected<br>insect<br>bite | Impetigo | Shingles | Total |
| --- | --- | --- | --- | --- | --- | --- | --- | --- |
| Feb-2024 | 41,010 | 32,678 | 18,893 | 18,384 | 3,412 | 6,815 | 4,078 | 125,270 |
| Mar-2024 | 54,753 | 34,535 | 22,888 | 17,220 | 4,180 | 6,739 | 4,069 | 144,384 |
| Apr-2024 | 56,924 | 41,951 | 19,295 | 17,714 | 6,099 | 7,191 | 4,470 | 153,644 |
| May-2024 | 58,021 | 44,339 | 20,239 | 15,909 | 19,010 | 6,235 | 4,691 | 168,444 |
| Jun-2024 | 52,288 | 43,289 | 15,826 | 15,052 | 27,294 | 6,362 | 4,626 | 164,737 |
| Jul-2024 | 54,859 | 51,321 | 17,682 | 14,115 | 36,370 | 6,740 | 5,103 | 186,190 |
| Aug-2024 | 40,771 | 53,389 | 17,915 | 10,970 | 44,391 | 7,448 | 5,151 | 180,035 |
| Sep-2024 | 49,991 | 56,235 | 15,680 | 15,059 | 25,348 | 8,390 | 4,867 | 175,570 |
| Oct-2024 | 64,617 | 62,297 | 19,971 | 22,456 | 14,715 | 9,394 | 5,344 | 198,794 |
| Nov-2024 | 72,132 | 61,395 | 28,205 | 26,379 | 7,846 | 10,254 | 5,175 | 211,386 |
| Dec-2024 | 97,961 | 65,292 | 37,957 | 37,330 | 5,074 | 10,179 | 5,530 | 259,323 |
| Jan-2025 | 93,311 | 60,923 | 26,038 | 38,113 | 4,330 | 9,967 | 5,257 | 237,939 |
| Total | 736,638 | 607,644 | 260,589 | 248,701 | 198,069 | 95,714 | 58,361 | 2,205,716 |
| Share | 33.4% | 27.5% | 11.8% | 11.3% | 9.0% | 4.3% | 2.6% |  |

**Figure A1:**
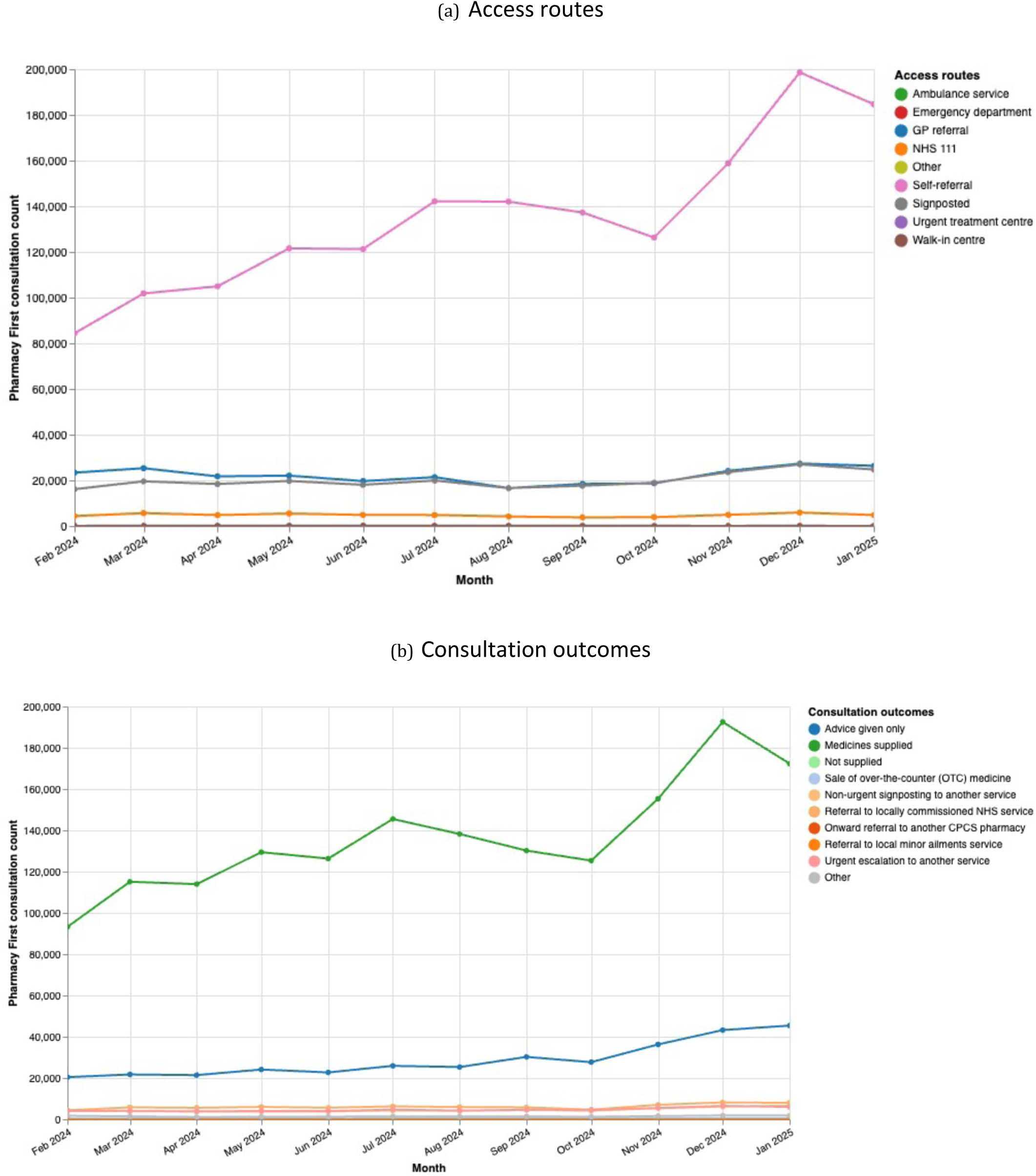
Monthly Pharmacy First consultation counts by (a) access route and (b) consultation outcome.

**Figure A2:**
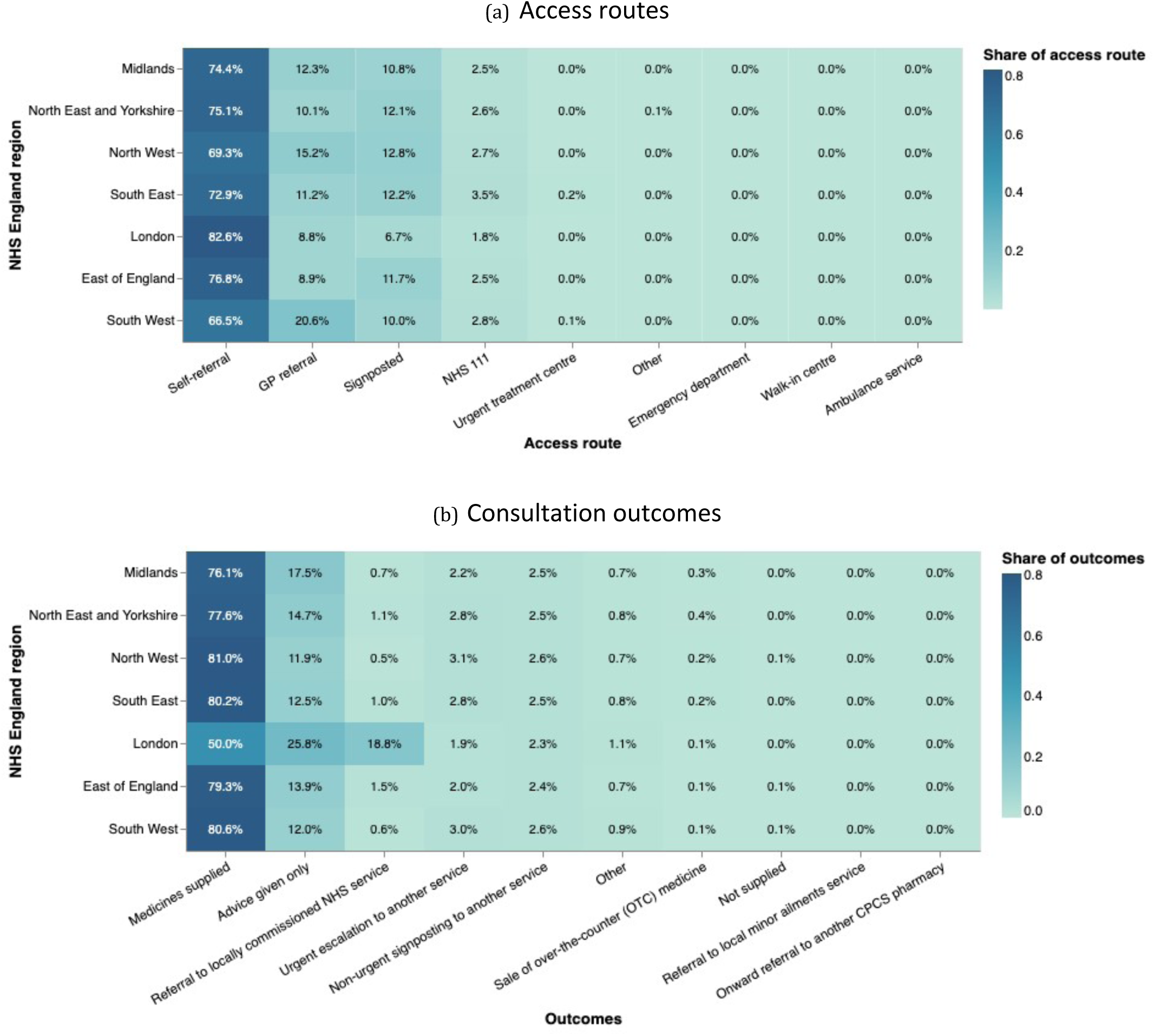
Distribution of Pharmacy First consultations by (a) access route and (b) consultation outcome at NHS England regional level. Within each NHS England region, percentages represent the proportion of consultations accounted for by each category shown in each subplot.

**Figure A3:**
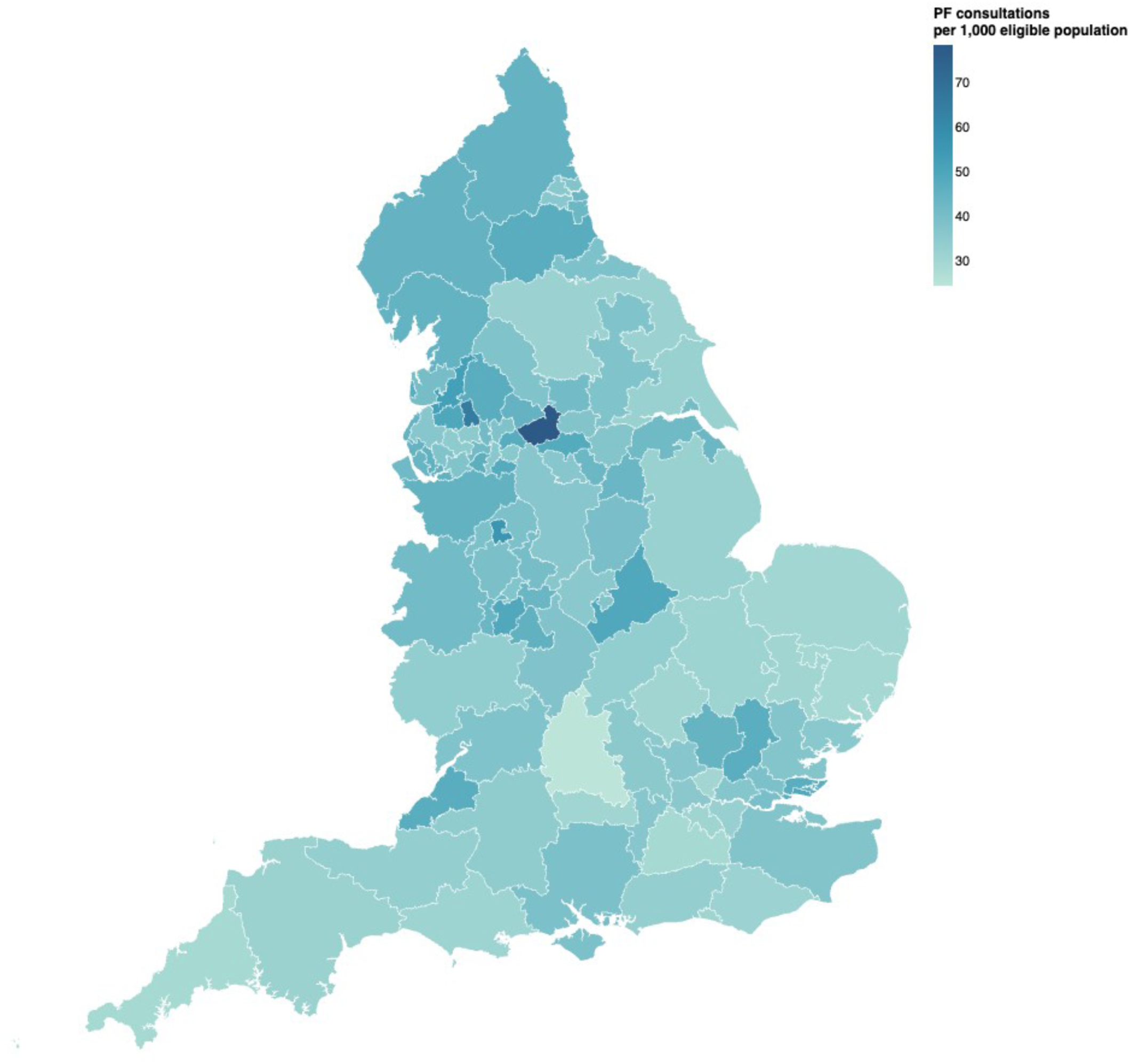
Distribution of Pharmacy First utilisation rates across Sub-ICBs, measured by consultations per 1,000 eligible population, ranging from 24.6 consultations per 1,000 eligible population in NHS Buckinghamshire, Oxfordshire and Berkshire West ICB 10Q to 78.7 in NHS West Yorkshire ICB X2C4Y.

**Figure A4:**
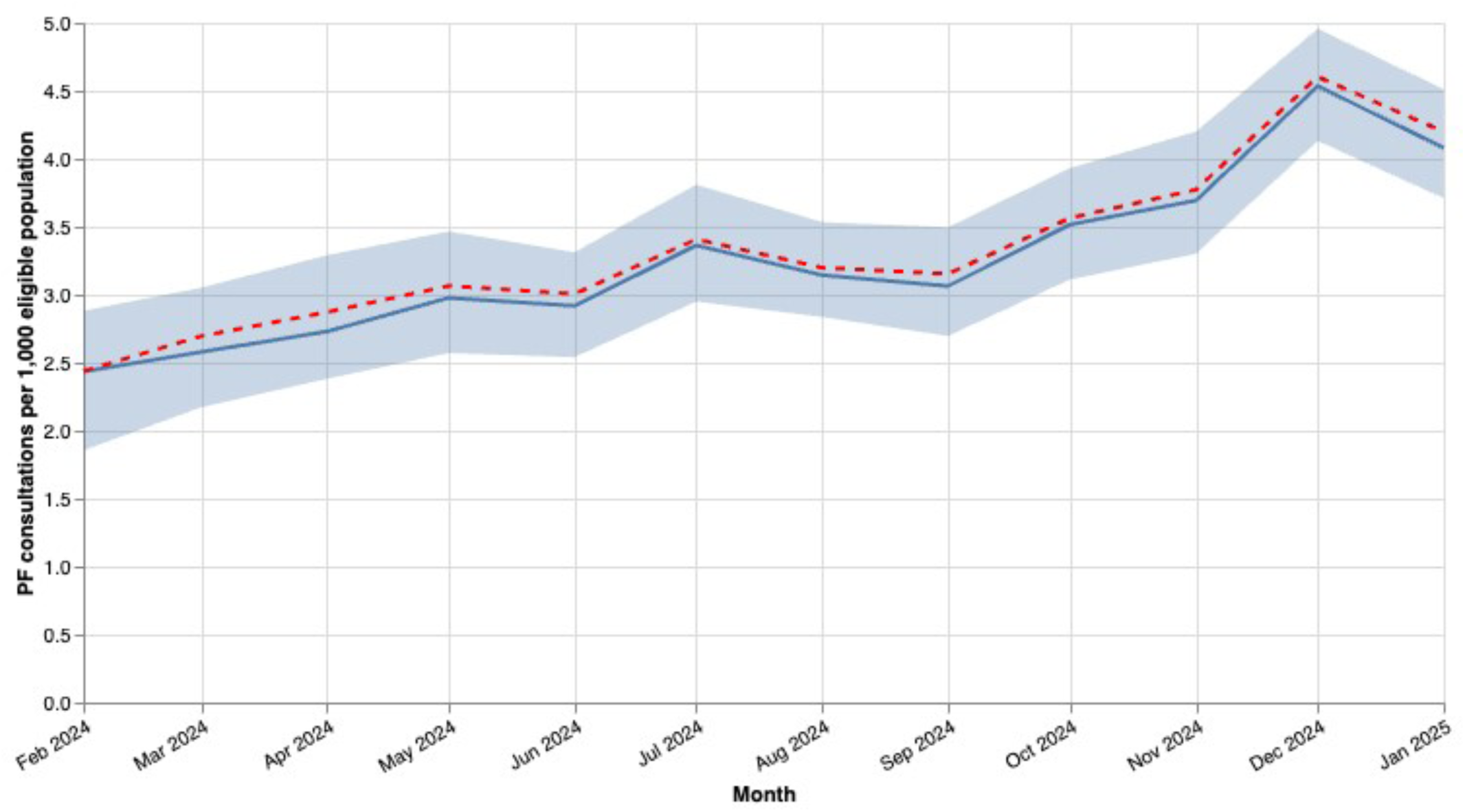
Monthly distribution of Pharmacy First utilisation rates across Sub-ICBs, measured by consultations per 1,000 eligible population. Median (solid line), mean (dashed line), and interquartile range (shaded area).

**Figure A5:**
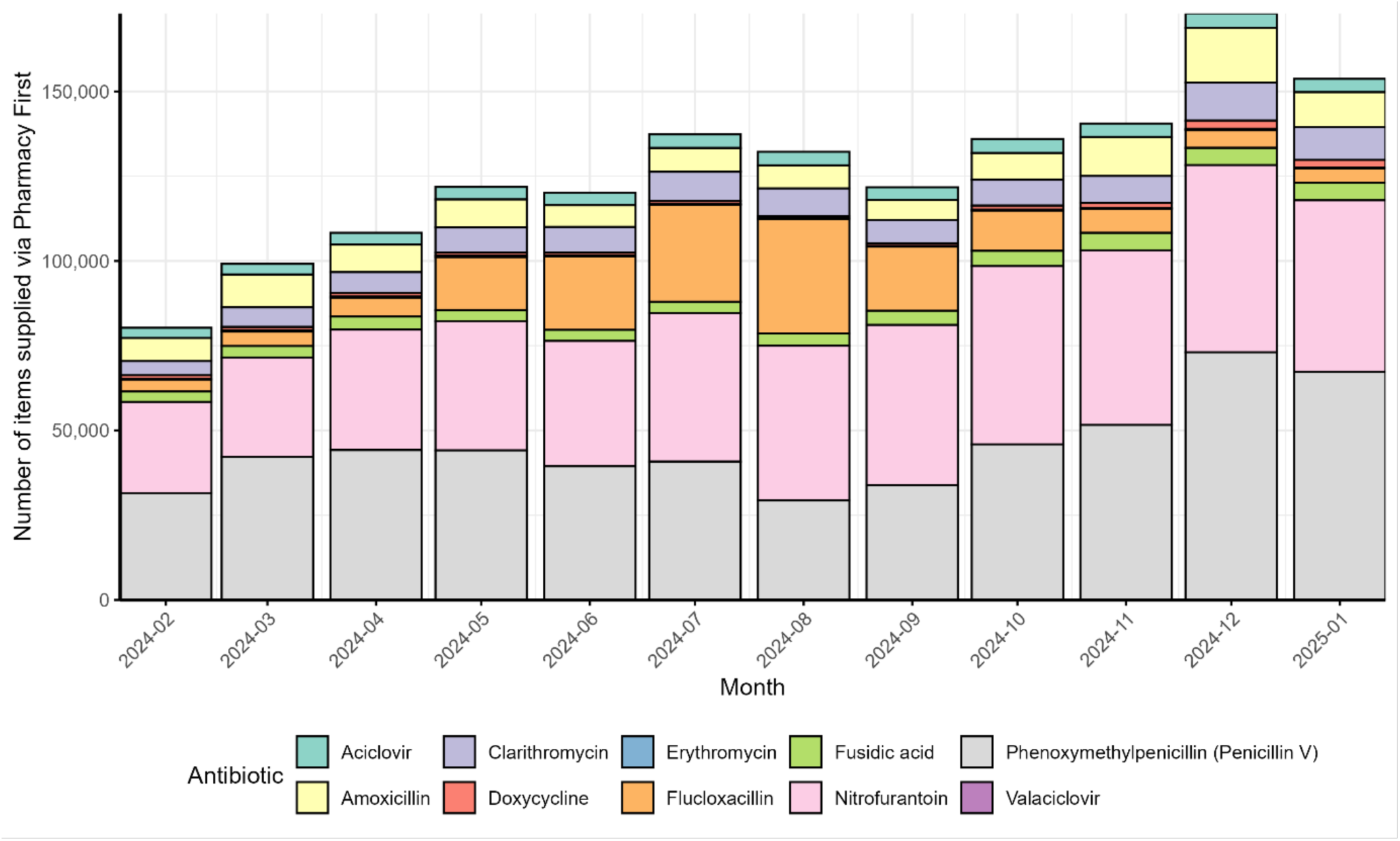
Total antimicrobial items supplied per month via Pharmacy First.

